# Protocol for ^18^F-PSMA PET imaging in staging and management of prostate cancer – a retrospective cohort study

**DOI:** 10.1101/2021.10.26.21265520

**Authors:** Matthew H V Byrne, Nithesh Ranasinha, Richard J Bryant, Freddie C Hamdy, Tom Leslie, Saiful Miah, Fergus Gleeson, Ruth MacPherson, Mark Tuthill, Andy Protheroe, Philip Camilleri, Phil Turner, Ami Sabharwal, Gerard Andrade, Alastair D Lamb

**Author notes:** **Corresponding author:** Name: Matthew H V Byrne, Email address, Full Institution address: Churchill Hospital, Old Road, Headington, Oxford, UK, Institution postcode: OX3 7LE. **Funding declarations:** None. **Study type:** Protocol. **Ethics:** This study was approved as a service evaluation by the Audit Department at Oxford University Hospitals Trust, UK and ethical approval was not required for this study.

## Abstract

**Background:** MRI, bone scan, and CT staging is recommended in the staging of prostate cancer. However, prostate-specific membrane antigen positron emission tomography (PSMA PET) could be superior in detection of local and distant prostate cancer cells. Most PSMA PET scans for prostate cancer are performed with a Gallium-68 ligand, with the Fluorine-18 (^18^F) ligand being introduced more recently. Methods: We will conduct a retrospective review of electronic patient records for all consecutive patients who underwent preoperative ^18^F-PSMA PET scan for prostate cancer from its introduction at our centre in 2019. We will compare PET scans with other imaging modalities and evaluate its use in diagnosis and management decisions for prostate cancer.

**Conclusions:** Understanding the role of ^18^F-PSMA PET in diagnosis and management could influence the diagnostic pathway of primary and secondary prostate cancer.

**Trial registration:** Not applicable.

## BACKGROUND

European Association of Urology guidelines recommend the use of magnetic resonance imaging (MRI) to assess local staging of prostate cancer alongside computer tomography (CT) and a bone scan to assess distant metastasis in patients with higher risk disease ^1^. However, MRI and CT have low sensitivity ^2,3^, and MRI has low specificity for detecting lymph node metastasis ^4–6^. This is problematic as patients considered appropriate for radical therapy for localised disease may have micro-metastases ^7^.

An alternative is positron emission tomography (PET), which can detect local and distant prostate cancer cells ^8^. ^18^F-fluorodeoxyglucose is a tracer that is used in most oncological PET scans ^9^, as it has preferential uptake in cancer cells ^10^. However, early studies into its use in prostate cancer were not promising, with one study showing a sensitivity of 37% ^11–13^, as such alternative PET tracers have been trialled ^9^. These include Gallium-68 (^68^Ga) and Fluorine-18 (^18^F) prostate-specific membrane antigen (PSMA) PET.

## ^68^Ga-PSMA-PET

^68^Ga-PSMA-PET is a ligand that binds to PSMA – a glycoprotein that is overexpressed on the surface of prostate cancer cells ^8^. In a randomised controlled trial of 302 men with high-risk prostate cancer, Hofman et al. evaluated the accuracy of ^68^Ga-PSMA-PET versus CT and bone scan at detecting pelvic nodal or distant metastatic disease. ^68^Ga-PSMA-PET had significantly superior accuracy (92% v 65%, p<0.0001), as well as higher sensitivity (85% v 38%) and specificity (98% v 91%) compared to CT and bone scan ^14^.

Petersen and Zacho conducted a systematic review of ^68^Ga-PSMA-PET for lymph node staging in prostate cancer. In 18 studies including 969 patients, the weighted sensitivity and weighted specificity compared to pathology as a reference was 59% (range 23-100%) and 93% (range 67-100%), respectively. In four studies, ^68^Ga-PSMA-PET was superior to CT or MRI, but in three studies comparing ^68^Ga-PSMA-PET to mpMRI (multiparametric MRI) or diffusion weighted MRI there were mixed results ^15^.

^68^Ga-PSMA-PET may also be used to improve the detection of metastatic disease in those with biochemical recurrence (BCR). In a systematic review of 37 studies comprising 4790 patients, Perera et al. demonstrated that in patients with biochemical recurrence (BCR) ^68^Ga-PSMA-PET positivity increased with increasing PSA level, and could be used to detect recurrence at low PSA levels (^68^Ga-PSMA-PET positivity for <0.2ng/ml was 33% and for 0,2-0.5ng/ml was 45%) ^16^.

Hofman et al. also found that first-line ^68^Ga-PSMA-PET changed management intent, modality, or modality delivery more commonly compared to CT and bone scan (28% v 15%, p=0.008). ^68^Ga-PSMA-PET was also associated with management change in those who received in second-line compared to CT and bonne scan (27% v 5%) ^14^. Han et al. performed a systematic review of the impact of ^68^Ga-PSMA-PET on management decisions. 1163 patients across 15 studies were included, and there were management changes in 54% (95% CI 47-60%) of patients following ^68^G-PSMA-PET ^17^. ^68^Ga-PSMA-PET positivity was significantly associated with increased rate of management change and meta-regression demonstrated that for each percent increase in positivity there was a 0.55% change in management (p<0.05).

## ^18^F-PSMA-PET

A more recent development in PSMA-PET imaging is the use of ^18^F-PSMA-PET as an alternative tracer to ^68^Ga, utilising the ^18^F radio-isotope ligand to label PSMA. One advantage of ^18^F is its minimal urinary excretion compared to ^68^G, which is limited by primary excretion through the urinary system, resulting in accumulation in the bladder, obscuring the prostate ^18^. Furthermore, ^18^F has practical advantages over ^68^Ga, including a longer half-life, aiding production of the agent in a central facility and distribution to distant imaging centres (Table 1) ^18^.

**Table 1:** Comparison of different PET modalities for prostate cancer imaging

| PET Modality | Mechanism of targeting |  | Sites of physiological uptake | Advantages | Disadvantages |
| --- | --- | --- | --- | --- | --- |
|  | Tracer | Target |  |  |  |
| **G68-PSMA-PET** | Gallium-68 radio-isotope | PSMA: transmembrane protein over-expressed in PCa | Kidney, bladder, salivary/lacrimal glands, liver, spleen, intestines | High prostatic specificity | Higher urinary clearance |
| **F18-PSMA-PET** | Fluorine-18 radio-isotope | PSMA: transmembrane protein over-expressed in PCa | Kidney, salivary/lacrimal glands, liver, spleen, intestines | High prostatic specificity; Minimal urinary clearance |  |

Prive et al compared ^18^F-PSMA-PET to conventional mp-MRI for local primary staging, in a cohort of 55 patients with intermediate to high risk prostate cancer^19^. 23 patients received both imaging modalities and underwent radical prostatectomy, with prostate specimens subsequently available for histopathological analysis. Using histopathological T stage as reference, this study demonstrated ^18^F-PSMA-PET correctly staged seminal vesical invasion (pT3b) more often than mp-MRI (90 vs 76%), whereas mp-MRI correctly staged extra-capsular extension (pT3a) more often than ^18^F-PSMA-PET (90 vs 57%).

Rowe et al evaluated ^18^F-PSMA-PET performance in distant lesion detection in metastatic prostate cancer^20^. Eight patients with biochemical recurrence underwent both ^18^F-PSMA-PET and CT-bone scan. 139 sites of PET positive for metastatic disease were detected, whereas only 45 sites of metastatic disease were identified on CT-bone scan. Although metastatic deposit biopsy data was not available for reference, regression analysis estimated 72% (95% CI 55-84%) of negative or equivocal lesions on CT-bone scan would be positive on ^18^F-PSMA-PET. Conversely, it was estimated that only 3% (95% CI 1-7%) of negative or equivocal lesions on ^18^F-PSMA-PET would be positive on CT-bone scan.

Dietlein et al compared ^18^F-PSMA-PET to ^68^Ga-PSMA-PET in the context of biochemically recurrent prostate cancer^21^. They performed PSMA-PET scans with both tracers on 25 patients with biochemically recurrent prostate cancer. This demonstrated non-inferiority of ^18^F-PSMA-PET and suggested its improved sensitivity in localising relapsed tumours after prostatectomy in moderately raised PSA contexts.

Our centre has performed ^18^F-PSMA PET since 2019 and we have conducted all primary staging with this modality since August 2020. In this study, we aim to examine the added value of ^18^F-PSMA PET over conventional staging modalities for primary and secondary disease, local and distant disease, and to investigate the influence of ^18^F-PSMA PET on treatment decisions.

### Hypothesis

^18^F-PSMA PET has suitable accuracy for investigation of primary and secondary prostate metastasis (nodal and distant) and alters management subsequently.

### Aims

- Comparison of PSMA-PET vs mpMRI for T and N stage
- Comparison of PSMA-PET vs MRI marrow, MRI Narrow Slice, or bone scan for M stage
- Comparison between imaging modalities for positive, negative, and equivocal results
- Comparison between imaging modalities and histology results
- Added value of PSMA-PET in clinically significant disease where there is a discordance between pathology findings and biochemistry
- Impact of PSMA-PET for primary staging on decision making and treatment strategy
- Impact of PSMA-PET for secondary staging (BCR post radical local therapy) on decision making and treatment strategy
- Influence of risk factors on the above aims – e.g. PSA level or D’Amico risk classification.

## METHODOLOGY

We will conduct a retrospective review of electronic patient records for all consecutive patients who underwent preoperative ^18^F-PSMA PET scan for prostate cancer from its introduction at the Churchill Hospital, Oxford, UK in 2019. Patients will be identified from the list of PET scans held by the radiology department and relevant data points will be collected (Table 2).

**Table 2:**
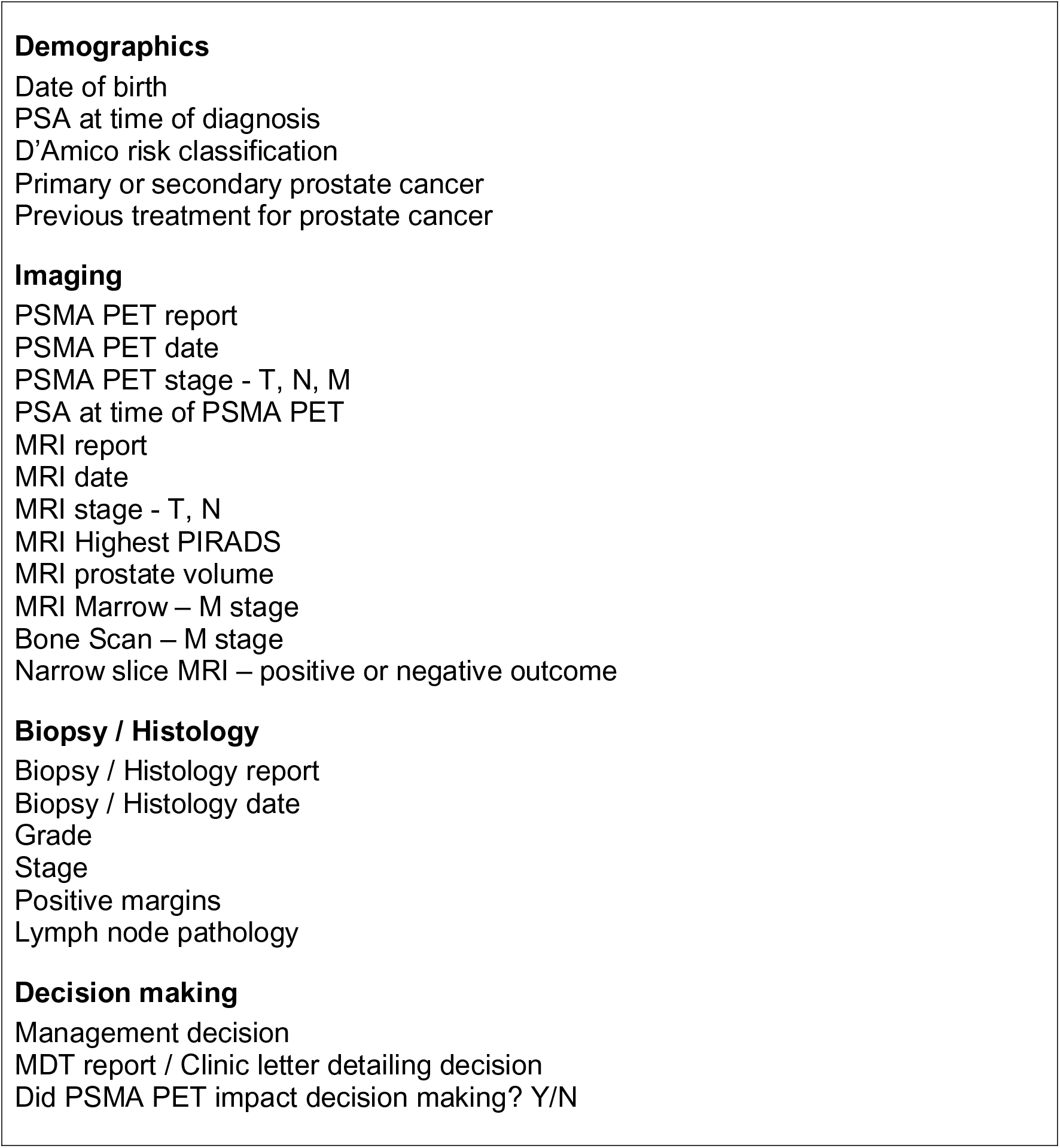
Data points which we intend to collect. As this study is a retrospective study, we may need to amend data points depending on whether the data is available

The STROBE (Strengthening the Reporting of Observational Studies in Epidemiology) guidelines for cohort studies will be followed ^22^.

## PICOS

Patients - all patients with prostate cancer who underwent a PSMA PET scan

Intervention - PET scan

Comparison - mpMRI, MRI Marrow, Bone scan, Pathological stage

Outcome - Diagnostic accuracy for stage & management implication

Study type - retrospective consecutive cohort study

## Data Availability

Not applicable.

## Notes

**Conflicts of interests:** None

### Competing Interest Statement

The authors have declared no competing interest.

### Funding Statement

This study did not receive any funding

### Author Declarations

This study was approved as a service evaluation by the Audit Department at Oxford University Hospitals Trust, UK and ethical approval was not required for this study.

## REFERENCES

1. Mottet N, van den Bergh RCN, Briers E, et al. EAU-EANM-ESTRO-ESUR-SIOG Guidelines on Prostate Cancer—2020 Update. Part 1: Screening, Diagnosis, and Local Treatment with Curative Intent. Eur Urol. 2021;79(2):243–262. doi:10.1016/j.eururo.2020.09.042

2. Hövels AM, Heesakkers RAM, Adang EM, et al. The diagnostic accuracy of CT and MRI in the staging of pelvic lymph nodes in patients with prostate cancer: a meta-analysis. Clin Radiol. 2008;63(4):387–395. doi:10.1016/j.crad.2007.05.022

3. Harisinghani MG, Barentsz J, Hahn PF, et al. Noninvasive Detection of Clinically Occult Lymph-Node Metastases in Prostate Cancer. N Engl J Med. 2003;348(25):2491–2499. doi:10.1056/nejmoa022749

4. Thoeny HC, Froehlich JM, Triantafyllou M, et al. Metastases in normal-sized pelvic lymph nodes: Detection with diffusion-weighted MR imaging. Radiology. 2014;273(1):125–135. doi:10.1148/radiol.14132921

5. Kiss B, Thoeny HC, Studer UE. Current Status of Lymph Node Imaging in Bladder and Prostate Cancer. Urology. 2016;96:1–7. doi:10.1016/j.urology.2016.02.014

6. Mottet N, van den Bergh RCN, Briers E, et al. EAU-ESTRO-ESUR-SIOG Guidelines on Prostate Cancer.; 2018. https://uroweb.org/wp-content/uploads/EAU-ESUR-ESTRO-SIOG-Guidelines-on-Prostate-Cancer-large-text-V2.pdf. Accessed June 11, 2021.

7. Sathianathen NJ, Geurts N, Nair R, Lawrentschuk N, Murphy DG, Lamb AD. The phytological future of prostate cancer staging: PSMA-PET and the dandelion theory. Futur Oncol. 2017;13(20):1801–1807. doi:10.2217/fon-2017-0074

8. Kiess AP, Banerjee SR, Mease RC, et al. Prostate-specific membrane antigen as a target for cancer imaging and therapy. Q J Nucl Med Mol Imaging. 2015;59(3):241–268. https://pubmed.ncbi.nlm.nih.gov/26213140/. Accessed June 11, 2021.

9. Fraum TJ, Ludwig DR, Kim EH, Schroeder P, Hope TA, Ippolito JE. Prostate cancer PET tracers: Essentials for the urologist. Can J Urol. 2018;25(4):9371–9383. https://pubmed.ncbi.nlm.nih.gov/30125515/. Accessed June 29, 2021.

10. Heiden MGV, Cantley LC, Thompson CB. Understanding the warburg effect: The metabolic requirements of cell proliferation. Science (80-). 2009;324(5930):1029–1033. doi:10.1126/science.1160809

11. Salminen E, Hogg A, Binns D, Frydenberg M, Hicks R. Investigations with FDG-PET scanning in prostate cancer show limited value for clinical practice. Acta Oncol (Madr). 2002;41(5):425–429. doi:10.1080/028418602320405005

12. Hofer C, Laubenbacher C, Block T, Breul J, Hartung R, Schwaiger M. Fluorine-18-fluorodeoxyglucose positron emission tomography is useless for the detection of local recurrence after radical prostatectomy. Eur Urol. 1999;36(1):31–35. doi:10.1159/000019923

13. Minamimoto R, Senda M, Jinnouchi S, et al. The current status of an FDG-PET cancer screening program in Japan, based on a 4-year (2006-2009) nationwide survey. Ann Nucl Med. 2013;27(1):46–57. doi:10.1007/s12149-012-0660-x

14. Hofman MS, Lawrentschuk N, Francis RJ, et al. Prostate-specific membrane antigen PET-CT in patients with high-risk prostate cancer before curative-intent surgery or radiotherapy (proPSMA): a prospective, randomised, multicentre study. Lancet. 2020;395(10231):1208–1216. doi:10.1016/S0140-6736(20)30314-7

15. Petersen LJ, Zacho HD. PSMA PET for primary lymph node staging of intermediate and high-risk prostate cancer: An expedited systematic review. Cancer Imaging. 2020;20(1). doi:10.1186/s40644-020-0290-9

16. Perera M, Papa N, Roberts M, et al. Gallium-68 Prostate-specific Membrane Antigen Positron Emission Tomography in Advanced Prostate Cancer— Updated Diagnostic Utility, Sensitivity, Specificity, and Distribution of Prostate-specific Membrane Antigen-avid Lesions: A Systematic Review and Meta-analysis. Eur Urol. 2020;77(4):403–417. doi:10.1016/j.eururo.2019.01.049

17. Han S, Woo S, Kim YJ, Suh CH. Impact of 68Ga-PSMA PET on the Management of Patients with Prostate Cancer: A Systematic Review and Meta-analysis. Eur Urol. 2018;74(2):179–190. doi:10.1016/j.eururo.2018.03.030

18. Giesel FL, Hadaschik B, Cardinale J, et al. F-18 labelled PSMA-1007: biodistribution, radiation dosimetry and histopathological validation of tumor lesions in prostate cancer patients. Eur J Nucl Med Mol Imaging. 2017;44(4):678–688. doi:10.1007/s00259-016-3573-4

19. Privé BM, Israël B, Schilham MGM, et al. Evaluating F-18-PSMA-1007-PET in primary prostate cancer and comparing it to multi-parametric MRI and histopathology. Prostate Cancer Prostatic Dis. 2021;24(2):423–430. doi:10.1038/s41391-020-00292-2

20. Rowe SP, Macura KJ, Mena E, et al. PSMA-Based [(18)F]DCFPyL PET/CT Is Superior to Conventional Imaging for Lesion Detection in Patients with Metastatic Prostate Cancer. Mol imaging Biol. 2016;18(3):411–419. doi:10.1007/s11307-016-0957-6

21. Dietlein F, Kobe C, Neubauer S, et al. PSA-Stratified Performance of (18)F- and (68)Ga-PSMA PET in Patients with Biochemical Recurrence of Prostate Cancer. J Nucl Med. 2017;58(6):947–952. doi:10.2967/jnumed.116.185538

22. von Elm E, Altman DG, Egger M, Pocock SJ, Gøtzsche PC, Vandenbroucke JP. The Strengthening the Reporting of Observational Studies in Epidemiology (STROBE) statement: guidelines for reporting observational studies. Lancet. 2007;370(9596):1453–1457. doi:10.1016/S0140-6736(07)61602-X

